# Gene fusions during the early evolution of mesothelioma correlate with impaired DNA repair and Hippo pathways

**DOI:** 10.1101/2023.04.20.23288867

**Authors:** Maymun Jama, Min Zhang, Charlotte Poile, Apostolos Nakas, Annabel Sharkey, Joanna Dzialo, Alan Dawson, Kudazyi Kutywayo, Dean A. Fennell, Edward J Hollox

## Abstract

Malignant pleural mesothelioma (MPM), a rare cancer a long latency period (up to 40 years) between asbestos exposure and disease presentation. The mechanisms coupling asbestos to recurrent somatic alterations are poorly defined. Gene-fusions arising through genomic instability may create novel drivers during early MPM evolution. We explored the gene fusions that occurred early in the evolutionary history. We conducted multiregional whole exome sequencing (WES) of 106 samples from 20 patients undergoing pleurectomy decortication and identified 24 clonal non-recurrent fusions, three of which were novel (*FMO9P-OR2W5*, *GBA3* and *SP9*). The number of early gene fusion events detected varied from zero to eight per tumour, and presence of gene fusions was associated with clonal SCNAs involving the Hippo pathway genes and homologous recombination DNA repair genes. Fusions involved known tumour suppressors *BAP1*, *MTAP*, and *LRP1B,* and a clonal oncogenic fusion involving *CACNA1D-ERC2, PARD3B*-*NT5DC2* and *STAB2*-*NT5DC2* fusions were also identified as clonal fusions. Gene fusions events occur early during MPM evolution. Individual fusions are rare as no recurrent truncal fusions event were found. This suggests the importance of early disruption of these pathways in generating genomic rearrangements resulting in potentially oncogenic gene fusions.

## Introduction

Malignant pleural mesothelioma (MPM) is a rare cancer that has increased in incidence over the past 5 decades across the world [1]. The United Kingdom has the highest MPM mortality rate in the world, with 6.25 per 100000 in men and 1.08 per 100000 in women [2], and is largely caused by exposure to asbestos dust [3].

Genomic analysis of MPM is important for two reasons. Firstly, it can help reveal the biology of tumour development during the years between asbestos exposure and presentation of disease. Secondly, it can identify key weaknesses in the tumour, for example loss of the *MTAP* gene encoding methylthioadenosine phosphorylase leads to an increase in the cellular concentration of substrate methylthioadenosine, which inhibits protein arginine methyltransferase 5 (PRMT5) which can be further inhibited by quinocrine [4-6]. Genomic analysis of somatic copy number alterations (SCNAs) and targeted sequencing in MPM initially identified *BAP1*, *CDKN2A/B*, and *NF2* as tumour suppressor genes frequently lost in MPM [7]. Subsequent whole exome and transcriptome analyses has increased the number of genes found to be recurrently mutated in MPM, with the implication that these genes are important driver loci where mutation drives the development of the tumour [8, 9].

Given the extensive heterogeneity within tumours, distinguishing individually rare driver loci from non-tumourogenic passenger loci can remain a challenge. One approach is based on the concept that a genomic alteration is more likely to be a driver event if it occurs early in tumour development. A multiregional approach, where tumour genomes are sampled from different physical regions of the same tumour, allows identification of genomic alterations occurring clonally, that is across all samples, and therefore likely to have occurred early in tumour development before differentiation in space and in genomic alteration content. In our MEDUSA multiregional exome study, we have determined the relative timing of somatic mutations and SCNAs in a series of tumours, and shown key early mutational events are common in MPM, including loss of chromosome 22q (which carries *NF2*) [10]. Furthermore, we have shown that that SCNA burden correlates with a high neutrophil:lymphocyte and platelet:lymphocyte ratio, both indicators of a systemic inflammatory response [11].

Gene fusions are an established mechanism of how structural variation in genomes can lead to a change in function, and many gene fusions are known to have an important role in cancer initiation and progression [12]. The well-known recurrent driver fusion *BCR-ABL1* in CML has shown to have a substantial impact on clinical and treatment outcome [13, 14]. The chimeric fusion codes for a protein that drives CML progression via aberrant tyrosine kinase activity of *ABL1* which signals and activates downstream oncogenic pathways [15]. *BCR-ABL1* was the first chimeric oncogenic protein that was drug targeted with a tyrosine kinase inhibitor [16]. Other examples include *IGH-MYC* predominantly seen in Burkitt’s lymphoma [17] and *TMPRSS2-ERG* and *TMPRSS2-ETV4* fusions in prostate cancers [18]. As well as providing information on the molecular mechanism of disease, and being potential drug targets, gene fusions can potentially encode tumour-specific antigens with the potential to be targets for cancer immunotherapy [19].

Although gene fusions are associated with activating oncogenes, most rearrangements reported so far in MPMs involve tumour suppressor genes and noncoding DNA [8, 20]. An analysis of the transcriptome of 216 MPMs along with matched normal samples using RNASeq found 43 recurrent gene fusions in 22 samples, confirmed using reverse transcription PCR [8]. These fusions mostly involved the tumour suppressor genes *NF2*, *BAP1, PTEN, PBRM1* and *SETD2*, and the function of the encoded proteins was likely to be disrupted. This study highlighted an alternative route for detecting tumour suppressor loss, for example the inactivation of *NF2* as a result of an inversion event generating *GSTT1-NF2* fusion. This alteration does not alter the gene dosage and would have been undetected without gene fusion analysis. Fusions involving *EWSR1* have been found in a subset of perotineal mesotheliomas, including *EWSR1-ATF1* [21] and *EWSR1-YY1* [22] but not in MPM. Gene fusion transcripts have been identified in large scale scans of RNASeq data from MPM tumours [23, 24], but the lack of timing information and, in most cases, the lack of matching non-tumour data, means the interpretation of these observations is unclear.

Identifying a gene fusion resulting from a SCNA in a tumour genome or fusion transcript from transcriptome data is not sufficient evidence to show that this is a driver alteration, indeed it is likely that most gene fusions are stochastic events and are passenger mutations [25] especially in tumours with a high burden of somatic structural variants. Here, we use multiregional sampling of individual tumours, combined with matched non-tumour samples, to identify gene fusions occurring early in the evolution of MPM. By using matched non-tumour exome data, multiple exomes of the same tumour, transcriptome data and PCR validation, we aimed to robustly identify early gene fusions more likely to be driver events that help drive MPM progression.

## Methods

### Ethical and governance approvals

The MEDUSA study was approved by NHS ethics committees under the reference 4/LO/1527 14/EM/1159.

### Tissue acquisition and processing

All patients had a confirmed histological diagnosis of MPM before recruitment into the MEDUSA cohort and were undergoing routine surgery involving extended pleurectomy decortication [10]. During surgery, samples were collected consistently from the same distinct sites of the tumour including (1) apex, (2) pericardium, (3) anterior costophrenic recess, (4) posterior costophrenic recess and (5) oblique fissure. Genomic DNA was extracted from peripheral blood and tumour tissue using a QiAamp mini kit (Qiagen). Total RNA from 100 μm sections of formalin-fixed, paraffin-embedded (FFPE) tissue was isolated using RNAprep pure tissue Kit from TIANGEN (DP431). RNA concentration was measured using Qubit RNA Assay Kit (Life Technologies, CA, USA) and RNA integrity was assessed using the Bioanalyzer 2100 system (Agilent Technologies, CA, USA).

### Whole exome sequencing

The GRCh37 reference genome was used throughout this project, and all genomic locations refer to that reference genome. Using the Agilent SureSelect Human All ExonV6 kit (Agilent), the WES library was prepared using 1 μg of genomic DNA derived from each tumour and matched peripheral blood samples. The genomic DNA was sheared into fragments of around 180-280bp using the hydrodynamic shearing system (Covaris) and fragments were then hybridised with biotinylated probes and captured by Streptomycin-coated magnetic beads to enrich Exonic regions via amplification. This is followed by paired end sequencing using the Illumina NovaSeq 6000 platform with a coverage of 276X at Novogene Ltd. After passing quality control, the reads were aligned to the genome assembly using Burrows-Wheeler Aligner (bwa-0.7.17). A combination of tools including Sambamba (v0.6.7) [26], Picard (v2.18.9), FastQC (v0.11.5) and SAMtools ref (v1.8) were used for data filtering and quality control.

### RNA sequencing

2 μg RNA per sample was used as input material for the RNA sample preparations. Sequencing libraries were generated using NEBNext® UltraTM RNA Library Prep Kit for Illumina® (NEB, USA) following manufacturer’s recommendations and index codes were added to attribute sequences to each sample. Briefly, mRNA was purified from total RNA using poly-T oligo-attached magnetic beads. After fragmentation, the first strand cDNA was synthesized using random hexamer primer followed by the second strand cDNA synthesis using dTTP. Remaining overhangs were converted into blunt ends via exonuclease/polymerase activities. After adenylation of 3’ ends of DNA fragments, NEBNext Adaptor with hairpin loop structure were ligated to prepare for hybridization. In order to select cDNA fragments of preferentially 150∼200 bp in length, the library fragments were purified with AMPure XP system (Beckman Coulter, Beverly, USA). The concentration of each library was measured with real-time PCR. Pools of the indexed library were then prepared for cluster generation and PE150 sequencing on Illumina NovaSeq 6000 at Novagene Ltd.

### Gene fusion detection

Meerkat detects gene fusions from the input aligned sequence file in BAM format by spanning each discordant read pair across the fusion partners [27]. With discordant read pair support, we can only indirectly infer that there is a breakpoint somewhere between the spanning regions. Split read mapping refines the breakpoint regions by local alignment. The structural variants (SVs) undergo a filtering pipeline to remove germline variants present in the reference and matched normal BAM files. A prediction was discarded if the fusion event is smaller than 100 bp or larger than 1Mb, or the fusion event is mapped to simple or satellite repeats, or the fusion event predicted has an extensive homology of more than 40 bp at the breakpoint junction, or the identity at the breakpoint is less than 20 bp for fusion events involving inter-chromosomal translocation.

Fusion events either contains more than 25% nonuniquely mapped reads, >3 discordant read pairs and >3 soft-clipped reads across the predicted fusion breakpoint in the normal matched genome were also discarded. Finally, intergenic-intergenic fusions, fusion events between paralogous genes, and fusions mapping to repetitive regions of the genome were removed, and truncal fusion events were selected. To confirm somatic status, fusion events were inspected using the Integrative Genome Browser (IGV).

Delly (version 0.8.7) [28] requires a joint input of tumour and matched normal sequencing data (BAM files), an indexed reference genome file and a list of genomic regions excluded from SV calling, such as centromeres and telomeres. The software predict SVs based on paired-end reads which can identify insert size distribution and read orientation, and split read support which provides single nucleotide resolution to characterise fusion breakpoints and investigate microhomologies or micro insertions. There are multiple filtering steps in between command runs in order to exclude any unmapped contigs and reads aligned to the centromere and telomere regions, as well as to remove any germline SVs detected in the matched normal exome. The output BCF (binary variant format) file containing the predicted somatic SVs, is converted to a VCF (variant calling format) file. To confirm somatic status, fusion events were inspected using the Integrative Genome Viewer (IGV) [29].

### Sanger sequencing

A duplex PCR assay was designed for each potential fusion, with one primer pair designed based on the chromosomal orientation of each fusion partner and the other primer pair amplifying an unrelated region of the human genome acting as a positive control. Primer sequences were constructed using Primer3 (version 0.4.0) and flanked a distance of 0.5 kb from the proximal and distal position of the breakpoint junction to ensure the PCR product spanned the predicted breakpoints for each fusion event. Following standard PCR (1.5mM MgCl_2_, Tris-ammonium sulphate buffer) and agarose gel electrophoresis, amplicons were extracted from the agarose gel using a Monarch® DNA Gel Extraction Kit (New England Biolabs) and Sanger sequenced using standard approaches (Eurofins Genomics (Cologne) and Source Biosciences (Nottingham)).

### Gene fusion detection from RNA sequencing

The STAR aligner (v.2.7.9a) [30] generates chimeric alignments from RNAseq data using a reference genome index file. The chimeric alignments are recorded in an BAM file, which is then used by Arriba (v.2.1.0) [31] to predict fusion events by searching for split and paired-end (discordant) reads. Fusion predictions were filtered for somatic fusions by discarding those detected in matched RNAseq data from whole blood. Next, likely recurrent artefacts were removed such as fusions predicted from read-throughs, non-canonical splicing or internal tandem duplication.

### Statistics

Comparison of SCNA counts in truncal gene fusion positive and negative tumours used a two-tailed Fisher’s exact test implemented in R v.4.1.0.

## Results

### Identification of truncal gene fusions

An overview of the approach used to detect gene fusions from exome data is shown in figure 1. Exomes from 20 patients were analysed using Meerkat, with 14 patients having exomes from four sampling sites and peripheral blood, and 6 patients (MED1, MED23, MED24, MED27, MED34, and MED37) with exomes from five sampling sites and peripheral blood. After filtering and visual inspection of the bam sequence alignments using Integrated Genomics Viewer (IGV), 23 truncal fusion events involving at least one gene were detected across five patients (table 1), with no truncal fusions seen in more than one patient (Supplementary table 2). For the 23 truncal fusion events detected, nine were a consequence of chromosomal translocations, six a consequence of inversions, five a consequence of duplications and three a consequence of deletions. Of the 23 truncal fusions identified using Meerkat, we found 21 by using Delly to independently identify SV breakpoints (table 1). In addition, another *SLC39A1* fusion (*ATP1A4*-*SLC39A1*) was also observed in patient 23, but only predicted in 3/5 regional tumour samples. An exception to the truncal rule (must be detected in 4-5 regional samples) was applied in this instance to further study the various SVs affecting the gene *SLC39A1* (table 1). To validate our gene fusions, we selected these 24 fusion events across different tumours, and 22 (92%) were confirmed by genomic PCR across the breakpoint and Sanger sequencing (table 1, Supplementary table 1), including the *ATP1A4*-*SLC39A1* fusion not being initially identified as truncal.

**Figure 1–.**
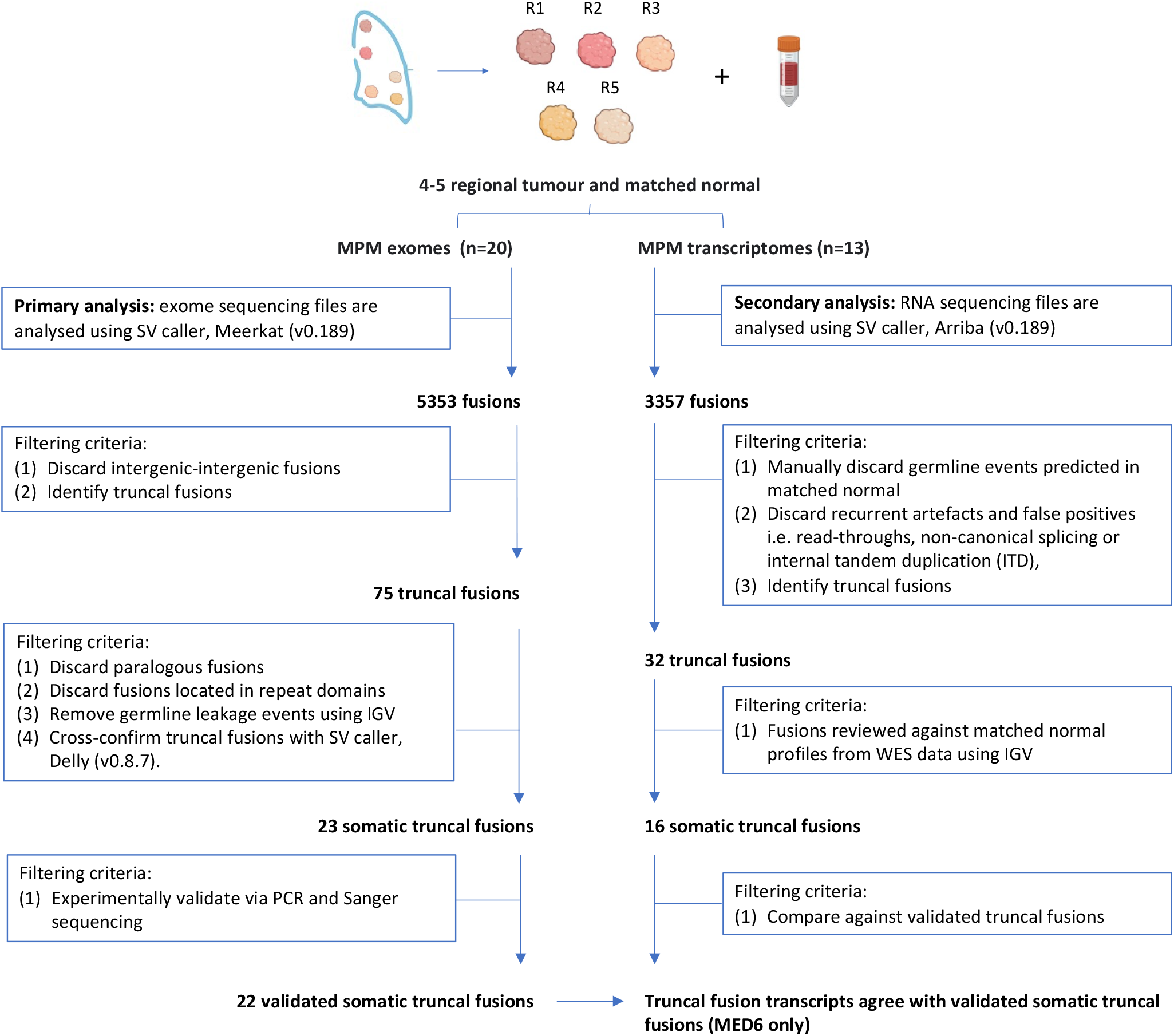
Overview of study

**Table 1-.** Truncal fusions identified in the MPM cohort from WES. The fusion breakpoints were predicted using Delly and Meerkat, using whole exome sequencing files as input. Different SVs generated the truncal fusion events such as inversions (INV), deletions (DEL), tandem duplications (DUP) and translocations (T). Truncal and branch events are represented as the “Tr” and “Br’, respectively, and ‘nd’ refers to as not detected. Fusion events were inspected using integrative genome browser (IGV) to confirm somatic (S) status. In the ‘PCR & Sanger’ column, positive and negative findings are indicated as ‘+’ and ‘–‘.

| Patient | Meerkat truncal fusion predictions | Breakpoint location | Chr | SV | Delly | IGV | PCR & Sanger |
| --- | --- | --- | --- | --- | --- | --- | --- |
| MED6 | igs - 5' *B3GALT2* | 1:193150615 – 1:207971545 | 1q | DUP | Tr | S | + |
| MED6 | igs - 5' *HSD17B4* | 5:118876469 – 5:118883187 | 5q | INV | Tr | S | + |
| MED6 | igs - 5' *HSD17B4* | 5:118814791 – 5:121213228 | 5q | DEL | Tr | S | + |
| MED6 | igs - 3' *AHRR* | 5:367885 – 5:16186887 | 5p | DUP | Tr | S | + |
| MED23 | *SLC39A1 – ATP1A2* | 1:153934727-160093945 | 1q | INV | Tr | S | + |
| MED23 | *ATP1A4 - SLC39A1 (branch)* | 1:160149525 – 1:153934766 | 1q | INV | Nd | S | + |
| MED23 | *EIF3J – SPG11* | 15:44834040-44865753 | 15q | DUP | Tr | S | + |
| MED23 | igs - 3' *IFNA22P* | 9:21260413-21278296 | 9p | INV | Tr | S | + |
| MED24 | *MTAP – GOLM1* | 9:21838005-88687558 | 9 | DEL | Tr | S | + |
| MED24 | igs - 3' *LRP1B* | 2:142354838-9:135566504 | 2q-9q | T | Tr | S | + |
| MED24 | igs - 3' *BAP1* | 3:52442553-3:88865856 | 3p | DEL | Tr | S | + |
| MED27 | *STAB1 – NT5DC2* | 3:52556557-52562845 | 3p | INV | Tr | S | + |
| MED27 | *PARD3B – NT5DC2* | 2:205655499-3:52568452 | 2q-3p | T | Tr | S | + |
| MED27 | *HRNR – LONP2* | 1:152185666-16:48358372 | 1q-16q | T | Tr | S | + |
| MED27 | *FMO9P – OR2W5* | 1:166575262-247654552 | 1q | DUP | Tr | S | + |
| MED27 | igs - 5' *GBA3* | 4:22749274-27435934 | 4p | INV | Br | S | + |
| MED27 | igs - 5' *EEF2* | 17:69159005-19:3976873 | 17q | T | Br | S | + |
| MED34 | *RNASEH2B – KHDRBS2* | 13:51503714-6:62736742 | 13q-6q | T | Tr | S | - |
| MED34 | *CACNA1D – ERC2* | 3:53535747-55937415 | 3p | INV | Tr | S | + |
| MED34 | *ARMC2 – C6orf170* | 6:109200964-121630288 | 6q | DUP | Tr | S | + |
| MED34 | igs - 5' *RALYL* | 6:143032099-8:85717151 | 6q-8q | T | Tr | S | + |
| MED34 | igs - 3' *SYDE2* | 1:85656316 – 18:57867181 | 1p-18q | T | Tr | S | + |
| MED34 | igs - 3' *SP9* | 2:175201394 – 9:116360081 | 2q-9q | T | Tr | S | + |
| MED27 | igs - 3' *STAB1* | 2:180151996-3:52553927 | 2q-3p | T | Tr | S | - |

To expand the scope of our analysis, we integrated publicly available data from a range of cancer types from CBioPortal (www.cbioportal.org). We extracted information on gene fusions, copy number alterations, and nonsynonymous mutations, and focused on our 22 validated fusion events from MPMs in this study. Twelve of our fusion events in MPMs had one fusion gene partner previously reported to be involved in a gene fusion in tumours, and seven fusions having both gene partners previously reported as a gene fusion in tumours (figure 2). The genes *HSD17B4, EEF2, TBC1D32, GOLM1*, and *PARD3B* have been reported as canonical fusions in tumours [32]. We found fusions involving *BAP1* and *MTAP* in patient MED24, with both fusions acting as a clonal second hit for these gene. Both *BAP1* and *MTAP* fusion genes have been previously reported in MPM, but none of the other fusions found had previously been reported in MPM. The majority of the genes involved in the fusions identified here (22/29) have been previously reported to be altered by copy number alterations or nonsynonymous mutations in MPM [9, 33], except *ARMC2, GOLM1, B3GALT2, OR2W5, LONP2, PARD3B*, and *SYDE2*, which may represent novel genes affected in MPM (figure 2). Only three of the genes involved in these fusions have been previously identified by Cosmic (v95) as being tumour suppressor genes (*BAP1*) or oncogenes (*CACNA1D*, *LRP1B*), further suggesting that the remaining fusions may highlight potentially novel genes involved in MPM.

**Figure 2–.**
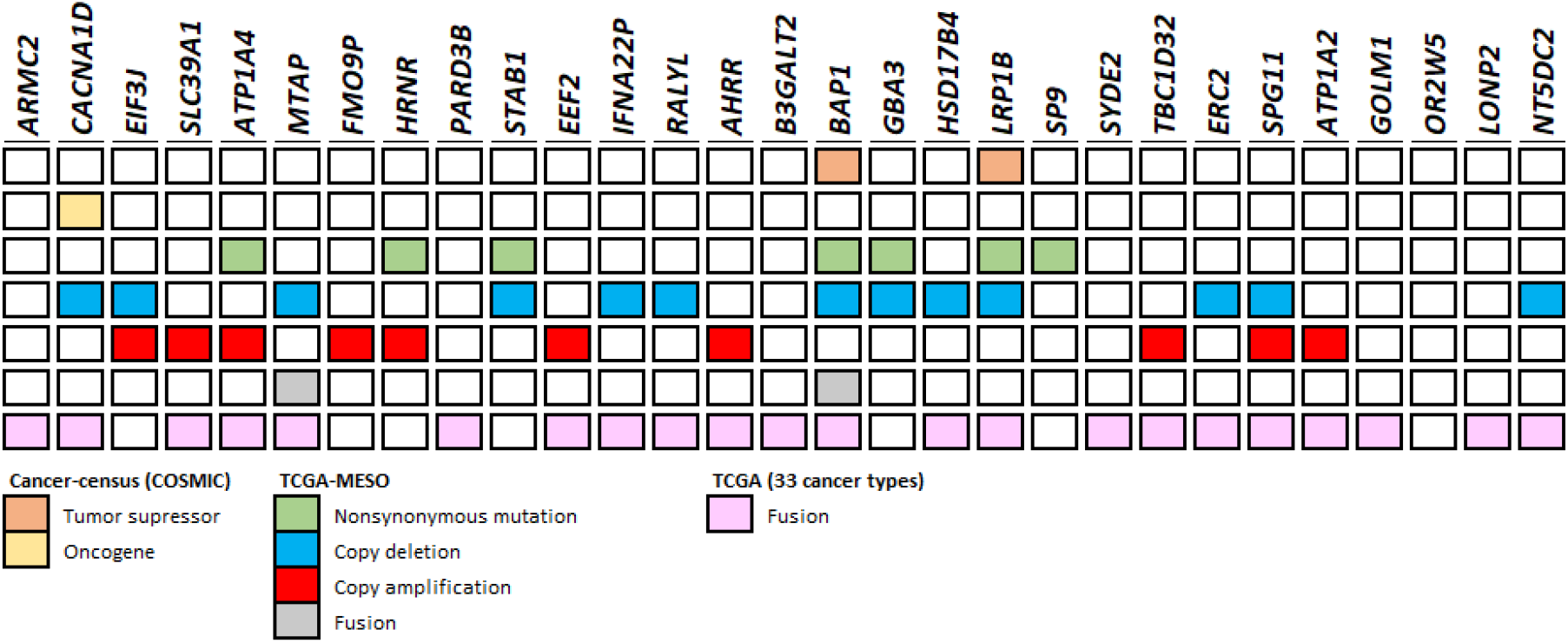
**Genes involved in the validated MPM truncal fusions.** For the genes affected by fusions in our cohort, the grid shows the extent of somatic alteration in MPM and across other tumour types as well as known cancer genes. Data from [9, 32, 33] and COSMIC v91

For 13 of the 20 patients, matched multiregional RNASeq data was available. A total of 950 fusion transcripts were identified in these 13 patients, of which 32 fusions were truncal, ranging from 0 to 8 fusions per patient. 16/32 were detected in corresponding WES profiles for each MPM using IGV (Supplementary table 3) and were also confirmed as somatic due to the absence of fusion event in the matched normal. Delly found all the truncal fusion predictions from the RNA seq data, in contrast to Meerkat, which detected 14 of the 16 with only 4 truncally (Table 2). Combining genomic and transcriptomic data, we found 36 tumour-specific novel truncal fusions across 9 patients (MED6, MED8, MED18, MED23, MED24, MED27, MED33, MED34, MED37) (Table 1 and Table 2).

**Table 2 -.** Truncal fusions identified in the MPM cohort from RNAseq. Different SVs generated the truncal fusion events such as inversions (INV), deletions (DEL), tandem duplications (DUP) and translocations (T). Truncal fusion calls from Arriba were compared to fusion data obtained from SV callers, Meerkat and Delly. Truncal and branch events were denoted as "Tr" and "Br," respectively, and "nd" indicated not detected.

| Patient | Arriba truncal fusion predictions | Breakpoint location | Chr | SV | Exome Seq |  |  |
| --- | --- | --- | --- | --- | --- | --- | --- |
|  |  |  |  |  | Meerkat | Delly | IGV |
| MED18 | *CNTNAP2-PTK2* | 7:146232929-8:141678452 | 7q-8q | T | Br | Tr | S |
| MED33 | *IFNA13-IFNA2* | 9:21367886-9:21385327 | 9p | INV | Br | Tr | S |
| MED37 | igs - 3' *BEND4* | 20:29565997-4:42122416 | 20q-4p | T | nd | Tr | S |
| MED37 | *FAM170A - RNF180* | 5:118970004-5:63645680 | 5q | INV | Br | Tr | S |
| MED37 | igs - 5' *ZNF746* | 7:149171891-11:34009480 | 7q-11p | T | Br | Tr | S |
| MED37 | igs - 5' *ZNF212* | 7:148936894-11:34009617 | 7q-11p | T | Br | Tr | S |
| MED37 | igs - 3' *OVCH2* | 11:7725478-11:8380941 | 11p | INV | Br | Tr | S |
| MED6 | *THSD4-UNC13C* | 15:72063526-15:54561803 | 15q | INV | Br | Tr | S |
| MED6 | *MYO9A - FBXL7* | 15:72144654-5:15855659 | 15q-5p | T | Br | Tr | S |
| MED6 | *GOLGA8B-CASC4* | 15:34825335-15:44677897 | 15q | INV | Br | Tr | S |
| MED6 | igs - 5' *SIRT7* | 17:79873586-17:79931695 | 17q | DUP | Br | Tr | S |
| MED8 | igs - 5' *HIF3A* | 19:46812453-19:40265152 | 19q | DUP | nd | Tr | S |
| MED6 | igs - 5' *B3GALT2* | 1:193150615 – 1:207971545 | 1q | DUP | Tr | Tr | S |
| MED6 | igs - 5' *HSD17B4* | 5:118876469 – 5:118883187 | 5q | INV | Tr | Tr | S |
| MED6 | igs - 5' *HSD17B4* | 5:118814791 – 5:121213228 | 5q | DEL | Tr | Tr | S |
| MED6 | igs - 3' *AHRR* | 5:367885 – 5:16186887 | 5p | DUP | Tr | Tr | S |

In our set of truncal fusions, the *SLC39A1*-*ATP1A2* and *ATP1A4*-*SLC39A1* fusion genes detected in patient MED23 are of particular interest. *SLC39A1* encodes a zinc transporter and is a known tumour suppressor gene controlled by YAP, part of the Hippo pathway [34]. The Hippo pathway is known to be dysregulated in mesothelioma and therefore loss of this gene is potentially a driver of mesothelioma [35]. Because the two fusion events breakpoints in *SLC39A1* are slightly different, and the two fusion events involve breakpoints in different members of the *ATP1A* family, two distinct inversions on both homologous chromosomes is possible. However, one event involving a single inversion followed by a deletion on one chromosome, is more likely (Figure 3).

**Figure 3–.**
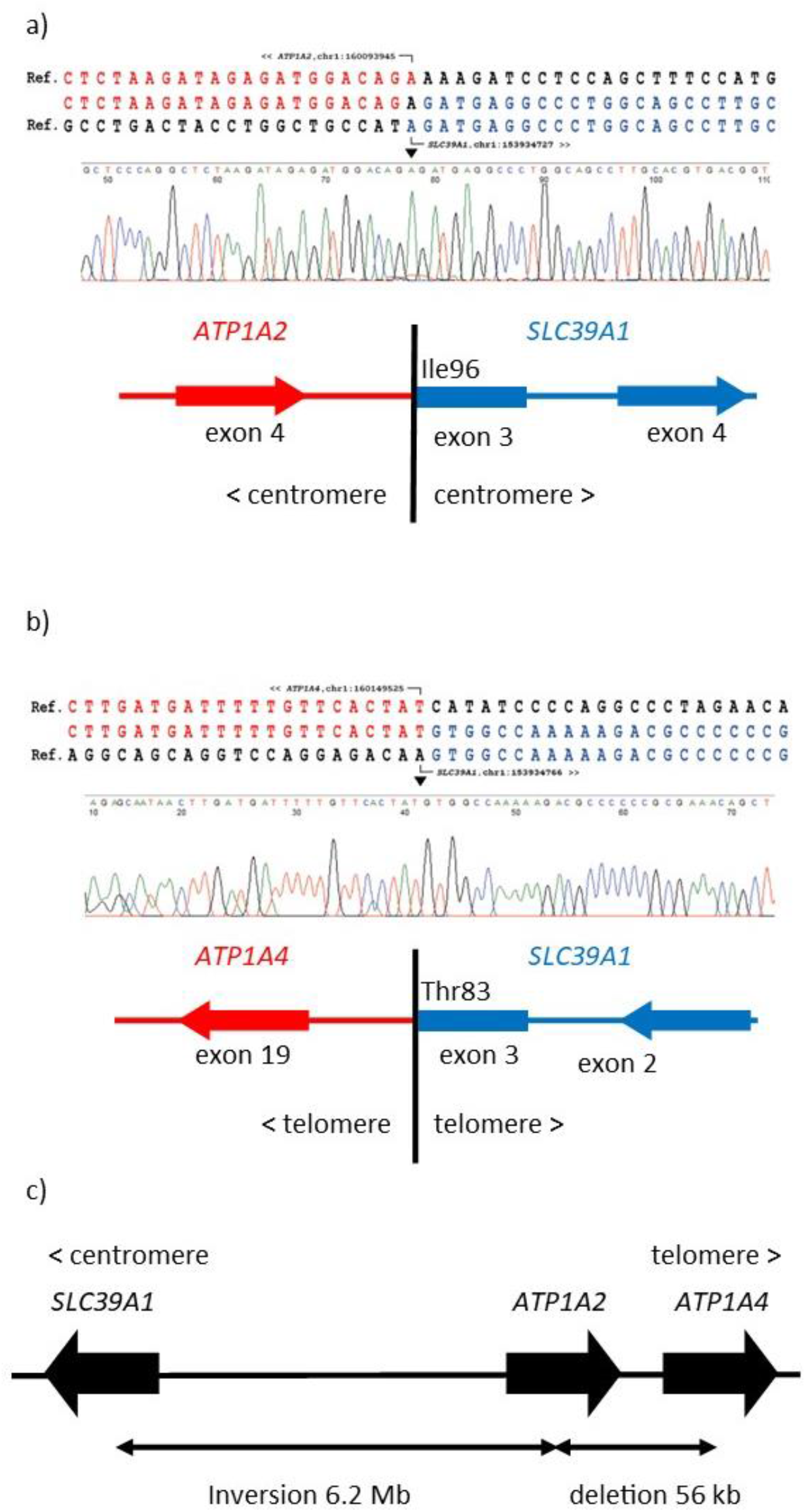
Validation of an inversion generating a *ATP1A2-SLC39A1* fusion gene a) Genomic Sanger sequence of breakpoint 1 of patient MED23 b) Genomic Sanger sequence of breakpoint 2 of patient MED23 c) Inferred structure of rearrangement based on breakpoint location and orienttation.

Although no gene fusions were recurrent across patients, patient MED6 showed distinct rearrangements affecting the *HSD17B4* gene on both alleles, implicating *HSD17B4* as a novel tumour suppressor gene in MPM (Figure 4). This interpretation was supported by transcript data which identified the truncated transcripts predicted by the exome analysis. The gene encodes the enzyme 17-beta-hydroxysteroid –dehydrogenase IV which is involved in fatty acid β-oxidation metabolism in peroxisomes and steroid metabolism [36, 37]. Its role in fatty acid metabolism potentially links it to feroptosis, a form of programmed cell death that plays a crucial role in tumour suppression.

**Figure 4–.**
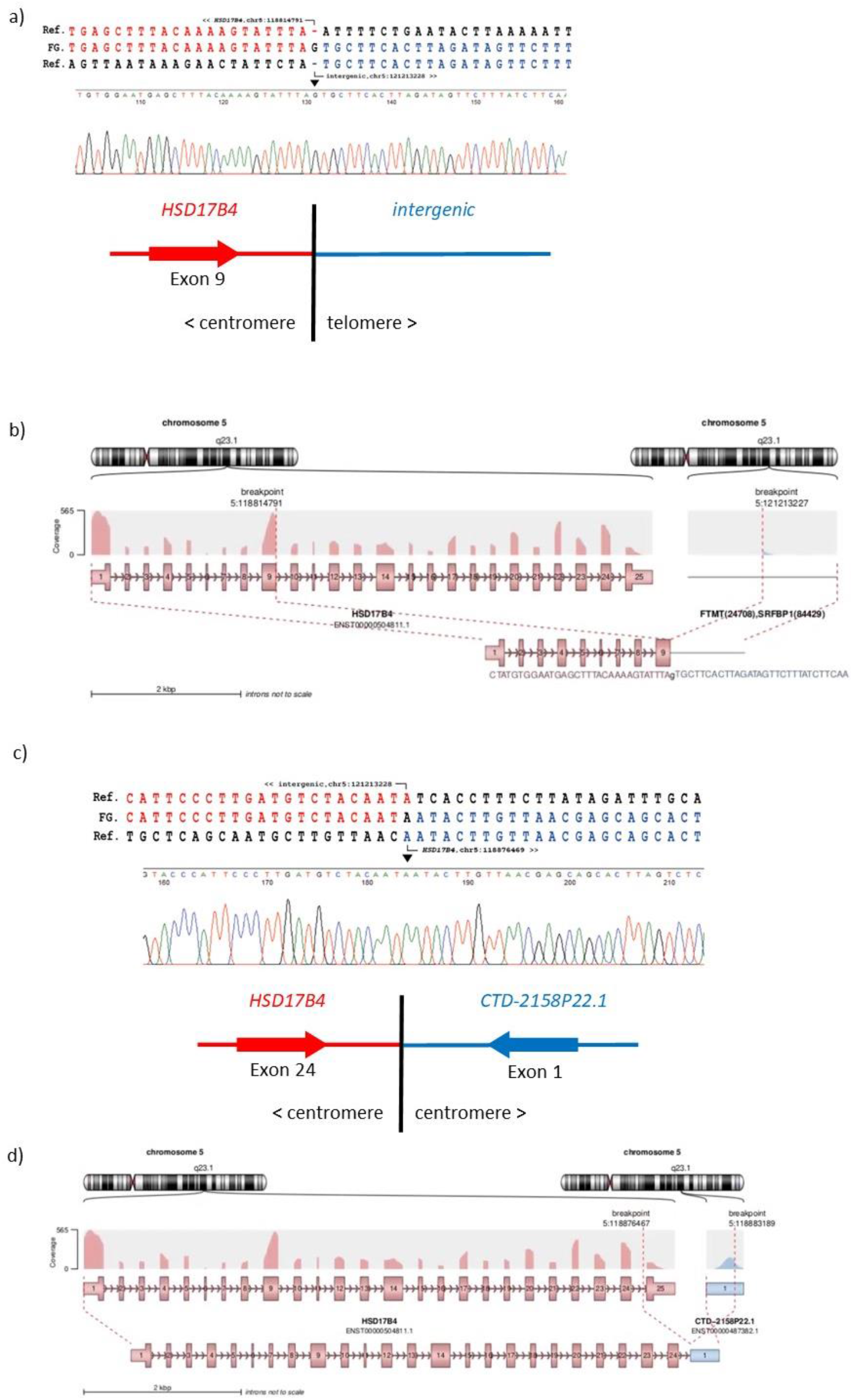
Validation of two fusion events disrupting *HSD17B4* a) Genomic Sanger sequence of *HSD17B4* inversion breakpoint 1 in patient MED6 b) *HSD17B4* inversion breakpoint allele 1 inferred from RNASeq data c) Genomic Sanger sequence of *HSD17B4* inversion breakpoint 1 in patient MED6 d) *HSD17B4* deletion breakpoint allele 2 inferred from RNASeq data

### Mechanism of gene fusion formation

Analysis of the gene fusion breakpoint at single nucleotide resolution allows assignment of likely mutational mechanisms that cause the structural variants that led to these gene fusions. Meerkat is able to automatically assign putative mechanism based on patterns of homology at the breakpoint inferred from the sequence alignment. Using the same decision tree that Meerkat uses, we were able to use the Sanger-verified breakpoint sequence and the breakpoint identified by Delly to assign putative mechanisms to each fusion (Supplementary table 4). All gene fusion events were predicted to have been caused by either alternative end joining or non-homologous end joining. For 27/36 truncal gene fusions, the mechanisms predicted by Delly and Meerkat agreed. For the 22 fusions with Sanger sequence across the breakpoint, we assigned 19 as being generated by NHEJ and 3 to alt-EJ (due to microhomology at the breakpoint), showing that early structural variation that generates gene fusions is driven by non-homologous end joining as a response to DNA double-strand breaks (Table 1, Supplementary table 4).

As the truncal gene fusions were found in a subset of patients analyzed (8 out of a total of 20 patients), we considered whether there was a particular molecular lesion or pattern correlated with, and possibly explaining, the presence or absence of truncal gene fusions in particular tumours. For example, *BAP1* mutation is a common driver of mesothelioma, and *BAP1* is a key component of the homologous recombination pathway, so we hypothesised that these 8 truncal fusion positive patients (MED6, MED23, MED24, MED27, MED34, MED18, MED33, and MED8) might all have mutations in *BAP1* and therefore a likely deficiency in homologous recombination (HR) repair, thereby promoting NHEJ-mediated repair of double-strand breaks. From the 14/20 patients where SCNA data were available from a previous study [10], we identified six of the eight truncal fusion positive tumours showed truncal *BAP1* loss from genomic analysis, in which 5/8 (MED24, MED18, MED33, MED27 and MED34) harboured a double-hit *BAP1* alteration. In comparison, only two of the six truncal fusion negative patients show *BAP1* loss, but this difference is not significant, possibly because of small numbers (p=0.2) (Supplementary table 5).

### Clonal fusions are assciated with impaired loss of homologous recombination DNA repair and Hippo pathway inactivation

In order to explore the basis of the difference between the truncal gene fusion-positive and truncal gene fusion-negative patients further, we compared truncal gene losses in the two groups to look for enrichment for particular pathways. This increases statistical power by analysing changes across multiple genes in a pathway, rather than individual genes. Using the gene list from the KEGG database (hsa03440), 33 out of 39 genes involved in HR were lost truncally, with enrichment in the 8 truncal fusion positive patients (p=0.01, two sided Fisher’s exact test) (Supplementary table 5). Analysis of the breakpoints suggests that this loss of HR gene function in accompanied by a switch to NHEJ (Supplementary table 4). Although clonal alterations do affect some NHEJ genes (KEGG hsa03450), there is no difference in the number of NHEJ genes affected between truncal gene fusion positive and negative patients (p=0.08, two sided Fisher’s exact test). We also analysed the Hippo pathway genes (KEGG hsa04390) in a similar manner and many more truncal alterations were found in truncal gene fusion positive patients than negative (p<0.0001, two sided Fisher’s exact test). Together this suggests that impaired Hippo pathway leading to DNA damage response [38] and impaired HR are responsible for an early, truncal gene fusion positive phenotype.

The above analysis is a correlation, and it could be argued that the high rate of gene fusions and high rate of gene loss are simply due to tumours from gene fusion positive tumours having a high number of SCNAs, as both gene fusions and gene loss are caused by SCNAs. However, there is no difference between gene-fusion-positive and gene-fusion-negative tumours in the proportion of the genome affected by truncal SCNAs (p=0.95, Wilcoxon rank sum test, data from [10]). Alternatively, the apparent enrichment of genes in particular pathways could be due to more genes of those pathways being present in the genome. If this was the case, however, we would expect there to be no relationship with time, in that the enrichment shown in later SCNA/SNV events would be the same as the enrichment shown in early events. Because we can distinguish early and late events in tumour evolution by clonal/subclonal analysis of the multiple regions of the tumour, we can test this by testing for enrichment when subclonal events are also included. For genes involved in HR and the Hippo pathway, this enrichment disappears when subclonal events are included, with no difference between truncal gene fusion positive patients and truncal gene fusion negative patients (Supplementary table 5). This strongly argues that early disruption of HR genes and Hippo pathway genes causes extensive further gene fusion events.

## Discussion

In this study we analysed exomes from multiple regions of malignant pleural mesothelioma in 20 patients to determine the extent and nature of gene fusion events that occurred early in the evolution of the tumour. Our results reveal the presence of *BAP1* and *MTAP* fusions, consistent with previous studies in the field (figure 2). However, we did not identify any potentially clonal truncal gene fusions involving the *NF2* gene, which has been reported in other studies (Bueno et al. 2016). Additionally, we did not observe any common recurrent gene fusions in our sample population. However, other clonal events merit further exploration, such as an in-frame *PARD3B*-*STAB1* fusion, as well as truncal biallelic alteration of *HSD17B4* encoding for an enzyme involved in steroid degradation in the peroxisome. These genes have been previously reported to be involved in tumour progression. We also identified a novel potentially oncogenic fusion (*CACNA1D*–*ERC2*) and an in-frame *PARD3B*-*NT5DC2* fusion in a subset of MPM samples using exome sequencing, although we were unable to validate the transcription of these fusions.

There are limitations to this study. Firstly, the number of patients is small so our power is limited to drawing only broad conclusions from our data. Extending patient cohorts and data sharing is critical particularly for rare tumour types such as MPM. Secondly, our analysis uses whole exome sequencing rather than whole genome sequencing, meaning that only a small proportion of the genome is sequenced at high coverage and many fusion events will be missed to due low coverage of sequence reads. Nevertheless, it is noticeable that at 200x coverage of the exome there are enough off-target sequence reads to identify at least some intergenic events, as 19 of 36 fusions identified involved a breakpoint in an intergenic region. These are potentially important, as they will inactivate the gene involved in the fusion.

Our key findings are that each patient had a unique constellation of truncal gene fusions, and truncal gene fusions occur only in a subset of MPMs, generating two classes of truncal gene fusion positive and negative tumours. For the gene fusion positive tumours, early truncal disruption by SCNAs are enriched in genes involved in the Hippo pathway and homologous repair genes. Our model is that, in some patients, early loss of key genes involved in DNA damage sensing and homologous repair causes a reliance on NHEJ and alt-EJ mechanisms to repair SV. These subsequent SVs can cause further loss of tumour suppressor genes or formation of oncogenic gene fusions. The overall burden of truncal SCNA does not differ between gene fusion positive and gene fusion negative tumours, so it is not clear why apparently similar levels SCNA should generate gene fusions in some tumours but not others. Our hypothesis is that this reflects the different sizes or different natures of SV alterations, so that truncal-gene fusion positive tumours either have smaller SCNAs, or more copy number neutral SVs, generated by NHEJ and alt-EJ mechanisms. A larger dataset with whole genome sequence is needed to reliably establish a more complete view of the SV landscape to test this idea.

Our results have clinical therapeutic implications. Because disruption of the Hippo pathway activates the YAP protein, small molecule inhibition of YAP has important therapeutic potential in MPM. Our results show that a subset of patients with high numbers of fusions may be especially disrupted in the Hippo pathway and therefore could be particularly responsive to such treatments. At present, NHEJ and alt-EJ inhibitors are currently in clinical and preclinical trails [39, 40], and gene fusion positive patients may show increased vulnerabilities to these inhibitors, allowing another approach to MPM treatment.

This study shows the benefit of multiregional sampling of tumours to detect genomic events occurring clonally across the tumour, and therefore implying that they occurred early in evolution. This has allowed us to identify potentially oncogenic gene fusions, and to construct a model of mesothelioma evolution for a subset of consistent with our data based on the early occurrence of genomic changes. The ability to deconstruct the timing of genomic events in a tumour has important therapeutic and diagnostic implications, particularly for mesothelioma, where the long latency and late diagnosis of the tumour remain a challenge for successful treatment of this cancer.

## Data Availability

Data for figure 5 and Supplementary table 5 are available at DOI: 10.25392/leicester.data.22360534
The exome sequencing data are available from the NCBI Sequence Read Archive under accession number PRJNA649889
All other data available from the corresponding author on request.

https://doi.org/10.25392/leicester.data.22360534

## Acknowledgements and funding

This work was supported by a British Lung Foundation-Mesothelioma UK grant MESOUK17-8. MJ was supported by a PhD studentship from the University of Leicester College of Life Sciences. This research used the ALICE High Performance Computing Facility at the University of Leicester.

## Author contributions

Funding acquisition: EJH and DF. Conceptualisation: EJH, MJ and DAF. Investigation: MJ, EJH, MZ. Resources: CP, AN, AS, JD, AD, KK and DAF. Writing – original draft: EJH and MJ. Writing – Review and Editing: all authors. Supervision: EJH and DAF.

## Data availability

Data for figure 5 and Supplementary table 5 are available at DOI: 10.25392/leicester.data.22360534

**Figure 5-.**
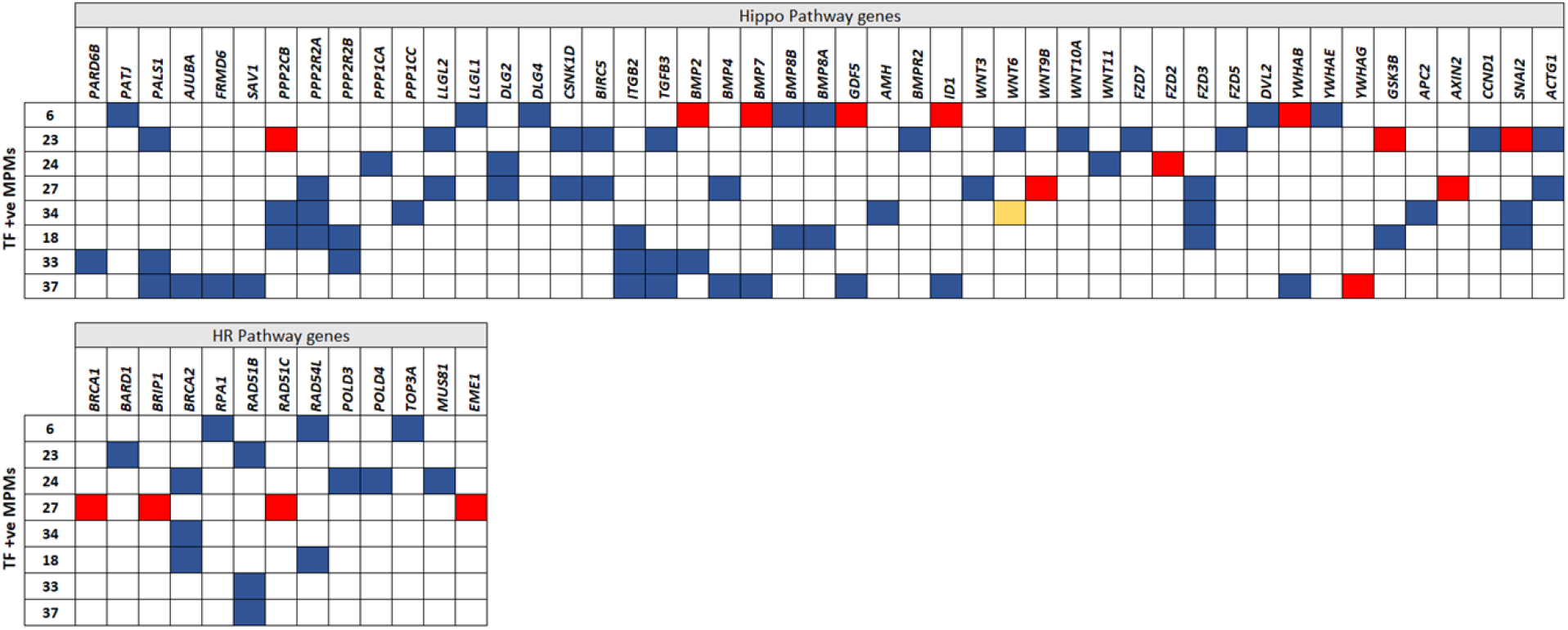
**Truncal alterations in HR and Hippo pathway genes enriched in truncal fusion positive MPMs.** Patients IDs are in rows and the genes in the partocular pathway are in columns. Truncal alterations are inferred if all the regional samples of a tumour have the same alteration, red denotes copy number gain, blue indicates copy number loss and yellow denotes somatic SNV. Source data from Zhang et al. (2021).

The exome sequencing data are available from the NCBI Sequence Read Archive under accession number PRJNA649889

All other data available from the corresponding author on request.

---

**Supplementary table 1:**
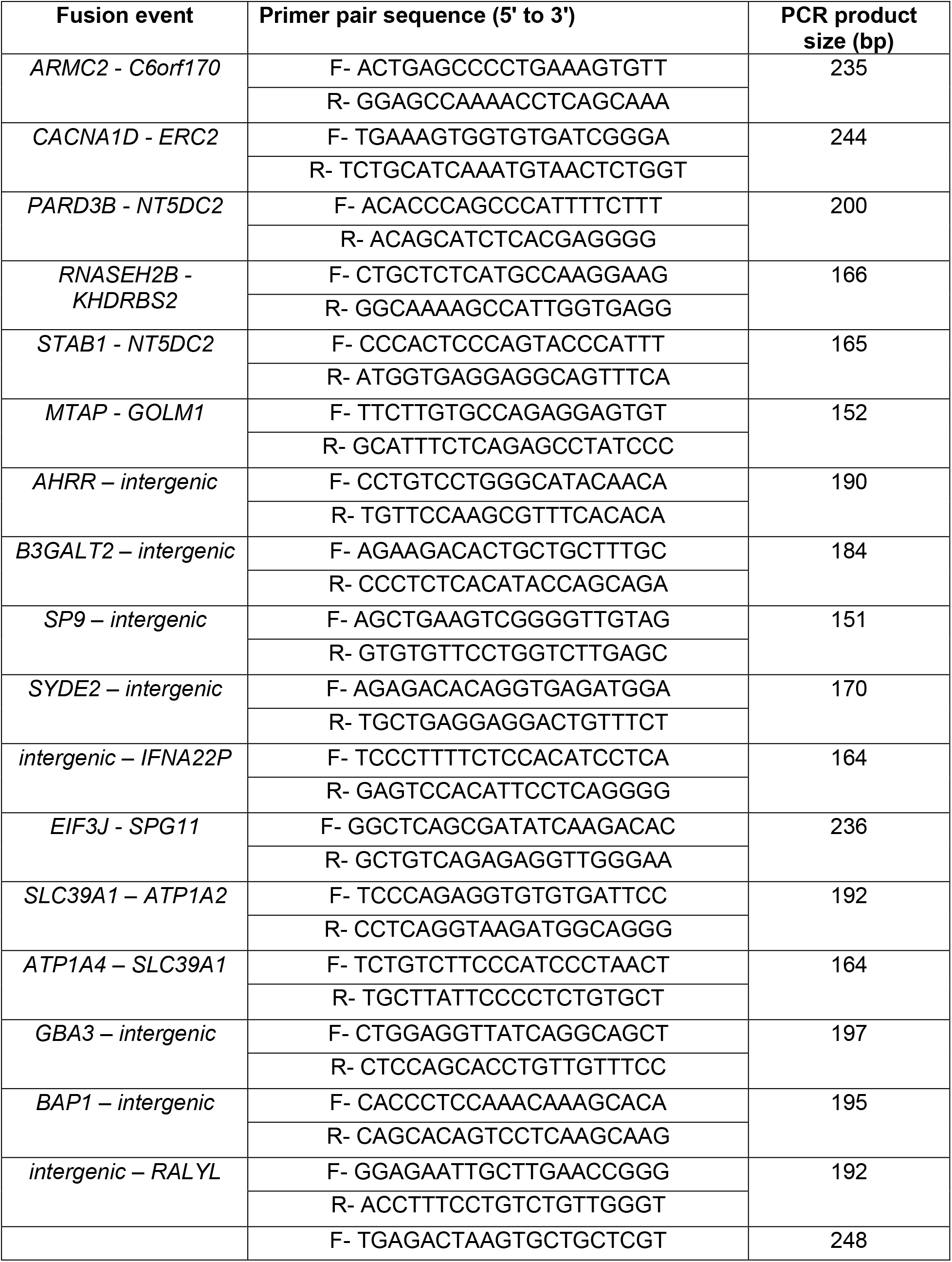
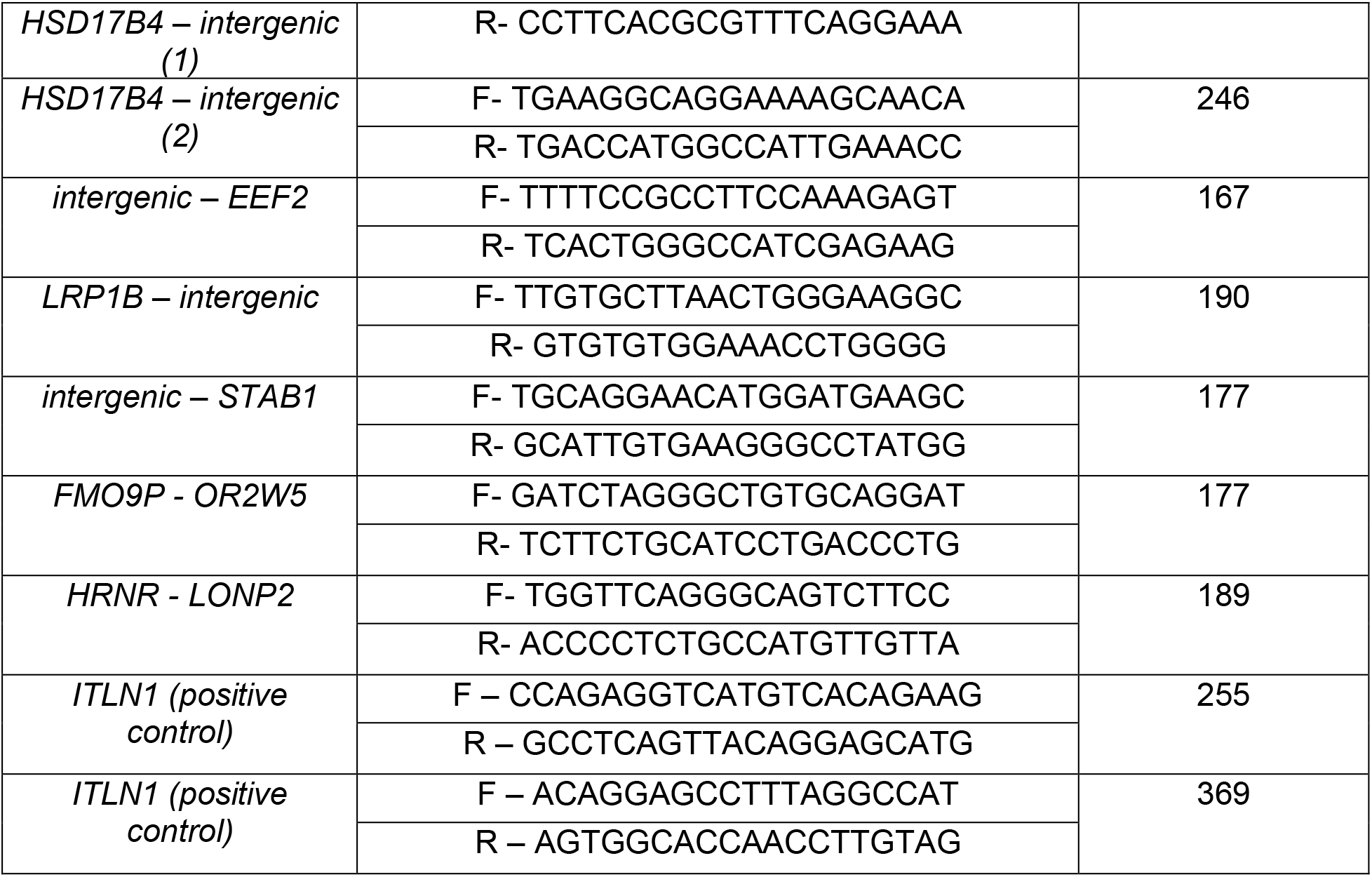
PCR primer sequences.

**Supplementary Table 2:** Number of fusions predictions identified per tumour from WES..

| Patient | Filtering |  |  | Truncal fusions |  |
| --- | --- | --- | --- | --- | --- |
|  | Pre- | Post- | Truncal fusions | Germline | Somatic |
| MED1 | 360 | 132 | 5 | 5 | 0 |
| MED3 | 131 | 27 | 0 | 0 | 0 |
| MED6 | 246 | 119 | 7 | 4 | 4 |
| MED7 | 173 | 81 | 3 | 3 | 0 |
| MED8 | 222 | 88 | 1 | 1 | 0 |
| MED9 | 291 | 101 | 7 | 7 | 0 |
| MED12 | 352 | 144 | 14 | 14 | 0 |
| MED16 | 222 | 96 | 3 | 3 | 0 |
| MED18 | 229 | 90 | 0 | 0 | 0 |
| MED20 | 245 | 93 | 5 | 5 | 0 |
| MED23 | 362 | 175 | 18 | 15 | 3 |
| MED24 | 347 | 173 | 16 | 13 | 3 |
| MED27 | 386 | 173 | 14 | 7 | 7 |
| MED32 | 254 | 94 | 2 | 2 | 0 |
| MED33 | 205 | 93 | 0 | 0 | 0 |
| MED34 | 368 | 167 | 21 | 15 | 6 |
| MED35 | 264 | 115 | 3 | 3 | 0 |
| MED37 | 235 | 79 | 0 | 0 | 0 |
| MED62 | 250 | 83 | 4 | 4 | 0 |
| MED64 | 211 | 84 | 5 | 5 | 0 |
| Total | 5353 | 2207 | 128 | 106 | 23 |

**Supplementary Table 3:** Number of fusions identified per tumour from RNAseq

| **Patient** | **Total fusions** | **Somatic fusions** | **After filtering** | **Truncal** | **WES** |
| --- | --- | --- | --- | --- | --- |
| MED6 | 284 | 94 | 78 | 8 | 8 |
| MED7 | 221 | 66 | 53 | 0 | 0 |
| MED8 | 241 | 77 | 63 | 3 | 1 |
| MED9 | 269 | 96 | 82 | 0 | 0 |
| MED16 | 269 | 104 | 83 | 0 | 0 |
| MED18 | 287 | 81 | 64 | 2 | 1 |
| MED20 | 256 | 94 | 78 | 0 | 0 |
| MED32 | 272 | 95 | 75 | 1 | 0 |
| MED33 | 245 | 103 | 81 | 5 | 1 |
| MED35 | 256 | 93 | 80 | 1 | 0 |
| MED37 | 292 | 146 | 110 | 6 | 5 |
| MED62 | 239 | 66 | 52 | 1 | 0 |
| MED64 | 226 | 62 | 51 | 1 | 0 |
| Total | 3357 | 1177 | 950 | 32 | 16 |

**Supplementary Table 4:** Sequence identity at the fusion breakpoint and the assigned double-strand break repair mechanism generating the truncal fusion events

| Fusion event | SV | Meerkat |  | Delly |  | Sanger |  |
| --- | --- | --- | --- | --- | --- | --- | --- |
|  |  | pSI (bp) | pSM | pSI (bp) | pSM | vSI (bp) | SM |
| intergenic - 5' *B3GALT2* | DUP | 3 | alt-EJ | 2 | NHEJ | 1 | NHEJ |
| intergenic - 5' *HSD17B4* | INV | 4 | alt-EJ | 4 | alt-EJ | 1 | NHEJ |
| intergenic - 5' *HSD17B4* | DEL | 1 (ins) | NHEJ | 6 | alt-EJ | 1 (ins) | NHEJ |
| intergenic - 3' *AHRR* | DUP | 4 | alt-EJ | 4 | alt-EJ | 2 (ins) | NHEJ |
| *SLC39A1* – *ATP1A2* | INV | 4 | alt-EJ | 4 | alt-EJ | 1 | NHEJ |
| *ATP1A4* - *SLC39A1* | INV | 0 | NHEJ | 1 | NHEJ | 0 | NHEJ |
| *EIF3J* – *SPG11* | DUP | 4 | alt-EJ | 7 | alt-EJ | 4 | alt-EJ |
| intergenic - 3' *IFNA22P* | INV | 1 | NHEJ | 2 | NHEJ | 1 | NHEJ |
| *MTAP* – *GOLM1* | DEL | 1 | NHEJ | 1 | NHEJ | 2 (ins) | NHEJ |
| intergenic - 3' *LRP1B* | T | 0 | NHEJ | 0 | NHEJ | 0 | NHEJ |
| intergenic - 3' *BAP1* | DEL | 1 (ins) | NHEJ | 2 | NHEJ | 1 (ins) | NHEJ |
| *STAB1* – *NT5DC2* | INV | 6 | alt-EJ | 6 | alt-EJ | 2 | NHEJ |
| *PARD3B* – *NT5DC2* | T | 3 | alt-EJ | 2 | NHEJ | 0 | NHEJ |
| *HRNR* – *LONP2* | T | 0 | NHEJ | 0 | NHEJ | 0 | NHEJ |
| *FMO9P* – *OR2W5* | DUP | 10 | alt-EJ | 7 | alt-EJ | 2 | NHEJ |
| intergenic - 5' *GBA3* | INV | 4 | alt-EJ | 4 | alt-EJ | 4 | alt-EJ |
| intergenic - 5' *EEF2* | T | 0 | NHEJ | 0 | NHEJ | 0 | NHEJ |
| *RNASEH2B* – *KHDRBS2* | T | 6 | alt-EJ | 5 | alt-EJ | - | - |
| *CACNA1D* – *ERC2* | INV | 4 | alt-EJ | 3 | alt-EJ | 1 | NHEJ |
| *ARMC2* – *C6orf170* | DUP | 2 | NHEJ | 4 | alt-EJ | 2 | NHEJ |
| intergenic - 5' *RALYL* | T | 0 | NHEJ | 4 | alt-EJ | 0 | NHEJ |
| intergenic - 3' *SYDE2* | T | 3 | alt-EJ | 5 | alt-EJ | 5 | alt-EJ |
| intergenic - 3' *SP9* | T | 1 | NHEJ | 1 | NHEJ | 1 | NHEJ |
| *CNTNAP2*-*PTK2* | T | 2 | NHEJ | 3 | alt-EJ | - | - |
| intergenic - 3' *STAB1* | T | 0 | NHEJ | 1 | NHEJ | - | - |
| *IFNA13*-*IFNA2* | INV | 2 | NHEJ | 3 | alt-EJ | - | - |
| intergenic - 3' *BEND4* | T | nd | - | 2 | NHEJ | - | - |
| *FAM170A* - *RNF180* | INV | 7 | alt-EJ | 3 | alt-EJ | - | - |
| intergenic - 5' *ZNF746* | T | 3 | alt-EJ | 3 | alt-EJ | - | - |
| intergenic - 5' *ZNF212* | T | 1 | NHEJ | 2 | NHEJ | - | - |
| intergenic - 3' *OVCH2* | INV | 0 | NHEJ | 2 | NHEJ | - | - |
| *THSD4*-*UNC13C* | INV | 6 | alt-EJ | 4 | alt-EJ | - | - |
| *MYO9A* - *FBXL7* | T | 3 | alt-EJ | 4 | alt-EJ | - | - |
| *GOLGA8B-CASC4* | INV | 1 | NHEJ | 1 | NHEJ | - | - |
| intergenic - 5' *SIRT7* | DUP | 4 | alt-EJ | 4 | alt-EJ | - | - |
| intergenic - 5' *HIF3A* | DUP | nd | - | 4 | alt-EJ | - | - |

**Supplementary Table 5:** Losses of genes in truncal fusion positive and negative tumours

| Genes | Truncal Fusions | Truncal loss | Non-Truncal or no loss | *p* | Truncal and Non-truncal losses | No loss | *p* |
| --- | --- | --- | --- | --- | --- | --- | --- |
| Hippo pathway | Positive | 245 | 835 | $5.3 \times 10^{-10}$ | 291 | 901 | 0.093 |
|  | Negative | 95 | 715 |  | 190 | 704 |  |
| HR pathway | Positive | 45 | 219 | 0.0018 | 57 | 247 | 0.49 |
|  | Negative | 14 | 184 |  | 37 | 191 |  |
| NHEJ pathway | Positive | 23 | 73 | 0.075 | 29 | 67 | 0.49 |
|  | Negative | 9 | 63 |  | 18 | 54 |  |
| *BAP1* | TF +ve | 6 | 2 | 0.2 | 6 | 2 | 0.5 |
|  | TF -ve | 2 | 4 |  | 3 | 3 |  |

## References

1 Zhai Z, Ruan J, Zheng Y, Xiang D, Li N, Hu J et al. Assessment of Global Trends in the Diagnosis of Mesothelioma From 1990 to 2017. JAMA Netw Open 2021; 4: e2120360.

2 Abdel-Rahman O. Global trends in mortality from malignant mesothelioma: Analysis of WHO mortality database (1994-2013). Clin Respir J 2018; 12: 2090–2100.

3 Liu B, van Gerwen M, Bonassi S, Taioli E, International Association for the Study of Lung Cancer Mesothelioma Task F. Epidemiology of Environmental Exposure and Malignant Mesothelioma. J Thorac Oncol 2017; 12: 1031–1045.

4 Mavrakis KJ, McDonald ER, 3rd, Schlabach MR, Billy E, Hoffman GR, deWeck A et al. Disordered methionine metabolism in MTAP/CDKN2A-deleted cancers leads to dependence on PRMT5. Science 2016; 351: 1208-1213.

5 Marjon K, Cameron MJ, Quang P, Clasquin MF, Mandley E, Kunii K et al. MTAP Deletions in Cancer Create Vulnerability to Targeting of the MAT2A/PRMT5/RIOK1 Axis. Cell Rep 2016; 15: 574–587.

6 Kryukov GV, Wilson FH, Ruth JR, Paulk J, Tsherniak A, Marlow SE et al. MTAP deletion confers enhanced dependency on the PRMT5 arginine methyltransferase in cancer cells. Science 2016; 351: 1214–1218.

7 Bott M, Brevet M, Taylor BS, Shimizu S, Ito T, Wang L et al. The nuclear deubiquitinase BAP1 is commonly inactivated by somatic mutations and 3p21.1 losses in malignant pleural mesothelioma. Nat Genet 2011; 43: 668–672.

8 Bueno R, Stawiski EW, Goldstein LD, Durinck S, De Rienzo A, Modrusan Z et al. Comprehensive genomic analysis of malignant pleural mesothelioma identifies recurrent mutations, gene fusions and splicing alterations. Nat Genet 2016; 48: 407–416.

9 Guo G, Chmielecki J, Goparaju C, Heguy A, Dolgalev I, Carbone M et al. Whole-exome sequencing reveals frequent genetic alterations in BAP1, NF2, CDKN2A, and CUL1 in malignant pleural mesothelioma. Cancer Res 2015; 75: 264-269.

10 Zhang M, Luo JL, Sun Q, Harber J, Dawson AG, Nakas A et al. Clonal architecture in mesothelioma is prognostic and shapes the tumour microenvironment. Nat Commun 2021; 12: 1751.

11 Templeton AJ, McNamara MG, Seruga B, Vera-Badillo FE, Aneja P, Ocana A et al. Prognostic role of neutrophil-to-lymphocyte ratio in solid tumors: a systematic review and meta-analysis. J Natl Cancer Inst 2014; 106: dju124.

12 Mertens F, Johansson B, Fioretos T, Mitelman F. The emerging complexity of gene fusions in cancer. Nat Rev Cancer 2015; 15: 371–381.

13 Stam K, Heisterkamp N, Grosveld G, de Klein A, Verma RS, Coleman M et al. Evidence of a new chimeric bcr/c-abl mRNA in patients with chronic myelocytic leukemia and the Philadelphia chromosome. N Engl J Med 1985; 313: 1429–1433.

14 Shtivelman E, Lifshitz B, Gale RP, Canaani E. Fused transcript of abl and bcr genes in chronic myelogenous leukaemia. Nature 1985; 315: 550–554.

15 Ren SY, Bolton E, Mohi MG, Morrione A, Neel BG, Skorski T. Phosphatidylinositol 3-kinase p85alpha subunit-dependent interaction with BCR/ABL-related fusion tyrosine kinases: molecular mechanisms and biological consequences. Mol Cell Biol 2005; 25: 8001–8008.

16 Druker BJ, Talpaz M, Resta DJ, Peng B, Buchdunger E, Ford JM et al. Efficacy and safety of a specific inhibitor of the BCR-ABL tyrosine kinase in chronic myeloid leukemia. N Engl J Med 2001; 344: 1031–1037.

17 Schmitz R, Ceribelli M, Pittaluga S, Wright G, Staudt LM. Oncogenic mechanisms in Burkitt lymphoma. Cold Spring Harb Perspect Med 2014; 4.

18 Cotter K, Rubin MA. The evolving landscape of prostate cancer somatic mutations. Prostate 2022; 82 Suppl 1: S13–S24.

19 Deininger MW, Druker BJ. Specific targeted therapy of chronic myelogenous leukemia with imatinib. Pharmacol Rev 2003; 55: 401–423.

20 Mansfield AS, Peikert T, Vasmatzis G. Chromosomal rearrangements and their neoantigenic potential in mesothelioma. Transl Lung Cancer Res 2020; 9: S92–S99.

21 Ren H, Rassekh SR, Lacson A, Lee CH, Dickson BC, Chung CT et al. Malignant Mesothelioma With EWSR1-ATF1 Fusion in Two Adolescent Male Patients. Pediatr Dev Pathol 2021; 24: 570–574.

22 Dermawan JK, Torrence D, Lee CH, Villafania L, Mullaney KA, DiNapoli S et al. EWSR1::YY1 fusion positive peritoneal epithelioid mesothelioma harbors mesothelioma epigenetic signature: Report of 3 cases in support of an emerging entity. Genes Chromosomes Cancer 2022; 61: 592–602.

23 Gao Q, Liang WW, Foltz SM, Mutharasu G, Jayasinghe RG, Cao S et al. Driver Fusions and Their Implications in the Development and Treatment of Human Cancers. Cell Rep 2018; 23: 227–238 e223.

24 Hu X, Wang Q, Tang M, Barthel F, Amin S, Yoshihara K et al. TumorFusions: an integrative resource for cancer-associated transcript fusions. Nucleic Acids Res 2018; 46: D1144–D1149.

25 Johansson B, Mertens F, Schyman T, Bjork J, Mandahl N, Mitelman F. Most gene fusions in cancer are stochastic events. Genes Chromosomes Cancer 2019; 58: 607–611.

26 Tarasov A, Vilella AJ, Cuppen E, Nijman IJ, Prins P. Sambamba: fast processing of NGS alignment formats. Bioinformatics 2015; 31: 2032–2034.

27 Yang L, Luquette LJ, Gehlenborg N, Xi R, Haseley PS, Hsieh CH et al. Diverse mechanisms of somatic structural variations in human cancer genomes. Cell 2013; 153: 919–929.

28 Rausch T, Zichner T, Schlattl A, Stutz AM, Benes V, Korbel JO. DELLY: structural variant discovery by integrated paired-end and split-read analysis. Bioinformatics 2012; 28: i333–i339.

29 Robinson JT, Thorvaldsdottir H, Wenger AM, Zehir A, Mesirov JP. Variant Review with the Integrative Genomics Viewer. Cancer Res 2017; 77: e31–e34.

30 Dobin A, Davis CA, Schlesinger F, Drenkow J, Zaleski C, Jha S et al. STAR: ultrafast universal RNA-seq aligner. Bioinformatics 2013; 29: 15–21.

31 Uhrig S, Ellermann J, Walther T, Burkhardt P, Frohlich M, Hutter B et al. Accurate and efficient detection of gene fusions from RNA sequencing data. Genome Res 2021; 31: 448–460.

32 Vellichirammal NN, Albahrani A, Banwait JK, Mishra NK, Li Y, Roychoudhury S et al. Pan-Cancer Analysis Reveals the Diverse Landscape of Novel Sense and Antisense Fusion Transcripts. Mol Ther Nucleic Acids 2020; 19: 1379–1398.

33 Hmeljak J, Sanchez-Vega F, Hoadley KA, Shih J, Stewart C, Heiman D et al. Integrative Molecular Characterization of Malignant Pleural Mesothelioma. Cancer Discov 2018; 8: 1548–1565.

34 Li L, Ugalde AP, Scheele C, Dieter SM, Nagel R, Ma J et al. A comprehensive enhancer screen identifies TRAM2 as a key and novel mediator of YAP oncogenesis. Genome Biol 2021; 22: 54.

35 Calvet L, Dos-Santos O, Spanakis E, Jean-Baptiste V, Le Bail JC, Buzy A et al. YAP1 is essential for malignant mesothelioma tumor maintenance. BMC Cancer 2022; 22: 639.

36 Amor DJ, Marsh AP, Storey E, Tankard R, Gillies G, Delatycki MB et al. Heterozygous mutations in HSD17B4 cause juvenile peroxisomal D-bifunctional protein deficiency. Neurol Genet 2016; 2: e114.

37 de Launoit Y, Adamski J. Unique multifunctional HSD17B4 gene product: 17beta-hydroxysteroid dehydrogenase 4 and D-3-hydroxyacyl-coenzyme A dehydrogenase/hydratase involved in Zellweger syndrome. J Mol Endocrinol 1999; 22: 227–240.

38 Pefani DE, O’Neill E. Hippo pathway and protection of genome stability in response to DNA damage. FEBS J 2016; 283: 1392–1403.

39 Kelm JM, Samarbakhsh A, Pillai A, VanderVere-Carozza PS, Aruri H, Pandey DS et al. Recent Advances in the Development of Non-PIKKs Targeting Small Molecule Inhibitors of DNA Double-Strand Break Repair. Front Oncol 2022; 12: 850883.

40 Li LY, Guan YD, Chen XS, Yang JM, Cheng Y. DNA Repair Pathways in Cancer Therapy and Resistance. Front Pharmacol 2020; 11: 629266.

